# United States Air Force Academy Class of 1967 Survival

**DOI:** 10.1101/2023.07.31.23293454

**Authors:** Harry Wetzler

## Abstract

**Introduction:** Air Force Academy cadets must meet medical standards to matriculate and be commissioned at graduation. However, during military service many graduates are exposed to significant occupational hazards. We examined the survival experience of the Air Force Academy class of 1967 (AFA67). Projections are provided for life expectancy and remaining quality adjusted life years, the number that will die in the next five years, and the number who will reach their 100^th^ birthday.

**Methods:** The dates of death as of September 1, 2022, were obtained from the Class of 1967 Memorial Ceremony booklet dated September 23, 2022. Birthdates were obtained for 94% of the known decedents. Simulated birthdates for the remainder were generated using a skew normal distribution. Likely death underreporting was accounted for. We used the National Health Interview Survey to derive US estimates to compare to AFA67. Parametric survival models using the Weibull distribution were used to calculate average survival time at age 78 (the median current age of AFA67), and standard life table techniques were used to estimate the number dying in the next five years and the number who will live to be centenarians.

**Results:** As of September 1, 2022, there were 130 known decedents, a 25.2% death incidence. After nearly fivefold excess mortality during the war in Southeast Asia, the class’s experience closely parallels that of contemporaneous college graduates. Adding 12 deaths to compensate for possible unreported deaths does not significantly alter the results. The point estimates for average life expectancy at 78, the number that will die in the next five years, and the number living to age 100 are 13.8 years, 74, and 76 respectively.

**Conclusion:** The AFA67 survival experience closely matches that of contemporaneous non-Hispanic white male college graduates in the US who were in excellent or very good health in their early twenties. There are numerous desirable opportunities to extend this research.

## INTRODUCTION

Humans in their 20s often have illusions of immortality, but as they reach their fifth or sixth decade, they begin to ask questions. How long will I live? How will I feel? What will I be able to do? How many of my friends will still be alive? The authors are members of the United States Air Force Academy class of 1967 (AFA67). We sought answers to these questions by looking at our class’s survival experience and comparing it to that of non-Hispanic white males (NHWM) in the US.

The published associations between health, longevity, and military service are inconsistent. Based on analysis of Health and Retirement Study (HRS) data from 1992-2006, Wilmoth and colleagues stated that “Vietnam veterans are in poorer health at the mean age [66.2], but they experience less substantial agerelated health changes than men who served during previous wars.”^1^ It is important to recognize that not all members of AFA67 served in Vietnam. Using data from the 2010 Behavioral Risk Factor Surveillance Survey, a U.S. population-based survey, Hoerster, et al, found that “despite better healthcare access, veterans as a group had poorer health and functioning than civilians and National Guard/Reserve members on several indicators.”^2^ di Lego, Turra, and Cesar reported that Brazilian Air Force officers had higher life expectancies than average Brazilians in 2000.^3^ However, younger Brazilian pilots had a higher risk of dying on duty compared to other officers but experienced lower mortality rates from other causes at advanced ages.

Education has long been recognized as an important factor in assessing survival and health. Sasson and Hayward reported that in 2017 the expected age at death for 25-year-old non-Hispanic white males (NHWM) with college degrees exceeded that of NHWM with a high school diploma or less education by 9.6 years.^4^ At age 75 or 80, the difference would be smaller, but these results illustrate the importance of considering education when analyzing longevity.

Poor health is associated with reduced life expectancy. Although there is no universally accepted measure of health, several studies have found associations between health status and subsequent mortality. DeSalvo, et al, found that those rating their health as ‘‘poor’’ had a 2-fold higher mortality risk compared with those rating their health as ‘‘excellent.’’^5^ Using more objective measures including the comprehensive metabolic profile, white blood cell differential count, and age, May and colleagues developed accurate prediction models for death after one year.^6^

This study had three objectives. First, analysis of deaths occurring in 1967-1973 during the Vietnam War. Second, deaths that occurred between 1974 and 2022 were compared to those of US NHWM college graduates. Finally, we projected the number dying in the next five years, the number living at age 100, and life expectancy at age 78 as well as life expectancies by level of self-reported health.

## METHODS

The 2007 Air Force Academy Register of Graduates published by the United States Air Force Academy Association of Graduates (AOG) was the source for denominator data.^7^ The names and dates of death for 129 class members were obtained from the Class of 1967 Memorial Ceremony booklet dated September 23, 2022, with data from the AOG. An additional death was identified after the booklet was published making a total of 130 known deaths as of September 1, 2022. Birthdates were obtained for 122 of the decedents and a convenience sample of 15 living graduates. Simulated birthdates for the remaining 379 (8 decedents without birthdates plus 371 presumed living) were generated using a skew normal distribution. Further details regarding birthdates are contained in the Supplement.

We used white male death rates from the 1967-1973 US life tables to calculate expected deaths during the war in Southeast Asia (SEA).^8^

Despite intense efforts to update the AFA67 death list prior to the publication of the Memorial Ceremony booklet in September 2022, 130 deaths are undoubtedly an undercount.^9^ To deal with this situation, we created an alternate cohort with 12 added deaths (hereafter referred to as AFA67+12) with ages at death approximately equal to those in the known deaths occurring after age 40.

Using simulated birthdate data for 379 of the graduates introduces error. To assess the impact of errors in birthdates, we looked at the relation between age at graduation and age at death as well as time to death for the 93 graduates with known birth- and death dates who died after 1973, i.e., after the conclusion of the SEA conflict. The results of these analyses are reported in Supplement.

For the AFA67 and AFA67+12 samples, we calculated ages at death and created life tables with censoring on September 1, 2022, for those assumed to be living. Standard US life tables do not account for education and health status. We used the National Health Interview Survey (NHIS), an annual survey of the US civilian non-institutionalized population (CNIP), to derive US estimates to compare to AFA67 and AFA67+12.^10^ Although NHIS respondents are interviewed only once, mortality follow-up based on the National Death Index (NDI) began in 1986. Thus, three groups were compared: AFA67, AFA67+12, and NHIS NHWM college graduates with years of birth 1943-1945 who were in excellent health when surveyed in 1986-1989.

To compare the three groups’ mortality and estimate life expectancies at age 78 along with the associated 95% margins of error, we fitted parametric survival models using the Weibull distribution for ages 43 and greater. Specifically, we used the streg module in Stata and estimated mean survival times. Since streg does not calculate other life table functions, we estimated the number dying in the next five years and the number living to age 100 by developing life tables using Gompertz modelled hazard rates and life expectancies at 78 forced to match the values from the Weibull parametric survival model. The 95% margins of error are the width of the 95% confidence intervals divided by 1.96.

We used NHIS survey data for years 2010-2018 with death follow-up through December 31, 2019, to estimate life expectancies at age 78 by self-reported health status. The analyses were confined to NHWM college graduates in the 76-80 age range at time of survey. The Fair and Poor health status categories were combined because there were very few respondents in the Poor category.

Empirical life tables, survival analyses, and parametric survival modelling were performed with Stata v 16. The NHIS analyses were weighted to better reflect the US CNIP. Gompertz based life tables were made in Excel.

## RESULTS

On June 7, 1967, 524 cadets graduated. Of these, two graduated posthumously, five were not commissioned, and one was a non-US cadet, leaving a total of 516 commissioned graduates. All graduates were males and virtually all were non-Hispanic whites. A total of 130 known deaths had occurred as of September 1, 2022. Table 1 is the life table for AFA67.

**Table 1.** Air Force Academy Class of 1967 Life Table as of September 1, 2022.

| Age | Alive | Deaths | Age | Alive | Deaths | Age | Alive | Deaths |
| --- | --- | --- | --- | --- | --- | --- | --- | --- |
| 22 | 516 | 1 | 41 | 481 | 1 | 60 | 456 | 1 |
| 23 | 515 | 2 | 42 | 480 | 0 | 61 | 455 | 6 |
| 24 | 513 | 11 | 43 | 480 | 4 | 62 | 449 | 3 |
| 25 | 502 | 1 | 44 | 476 | 0 | 63 | 446 | 4 |
| 26 | 501 | 10 | 45 | 476 | 0 | 64 | 442 | 1 |
| 27 | 491 | 1 | 46 | 476 | 0 | 65 | 441 | 2 |
| 28 | 490 | 3 | 47 | 476 | 0 | 66 | 439 | 1 |
| 29 | 487 | 1 | 48 | 476 | 1 | 67 | 438 | 2 |
| 30 | 486 | 0 | 49 | 475 | 3 | 68 | 436 | 3 |
| 31 | 486 | 0 | 50 | 472 | 3 | 69 | 433 | 8 |
| 32 | 486 | 0 | 51 | 469 | 0 | 70 | 425 | 1 |
| 33 | 486 | 1 | 52 | 469 | 3 | 71 | 424 | 5 |
| 34 | 485 | 0 | 53 | 466 | 1 | 72 | 419 | 3 |
| 35 | 485 | 1 | 54 | 465 | 1 | 73 | 416 | 4 |
| 36 | 484 | 1 | 55 | 464 | 1 | 74 | 412 | 7 |
| 37 | 483 | 1 | 56 | 463 | 2 | 75 | 405 | 7 |
| 38 | 482 | 1 | 57 | 461 | 1 | 76 | 398* | 8 |
| 39 | 481 | 0 | 58 | 460 | 1 | 77 | 338* | 3 |
| 40 | 481 | 0 | 59 | 459 | 3 | 78 | 101* | 1 |

\*censored (52, 234, and 81 withdrawn alive at ages 76, 77, and 78, respectively). Note that there are several ages where no deaths are known to have occurred.

### Excess deaths 1967-73 due to Southeast Asia conflict

In the life table, it is apparent that many deaths occurred at ages 24 through 28. When the class of 1967 graduated in June 1967, the pace of US activity in the SEA conflict was increasing. By the end of 1973, when the median age was 28, 19 class members had been killed in SEA and another 10 died from other causes including 4 because of aircraft accidents outside the combat zone. Thus, a total of 29 died by the end of 1973. During that period, only 6 deaths would have been expected based on the US white male death rates at that time.

#### Post-SEA conflict experience, 1974-2022, with comparison to comparable US population

After one death in 1974, there were no known deaths until June 1978 when the decedent was 33 years old. Due to the relatively small sample, there is considerable variation in deaths by age as seen in the chart below. However, the number of deaths increases with age.

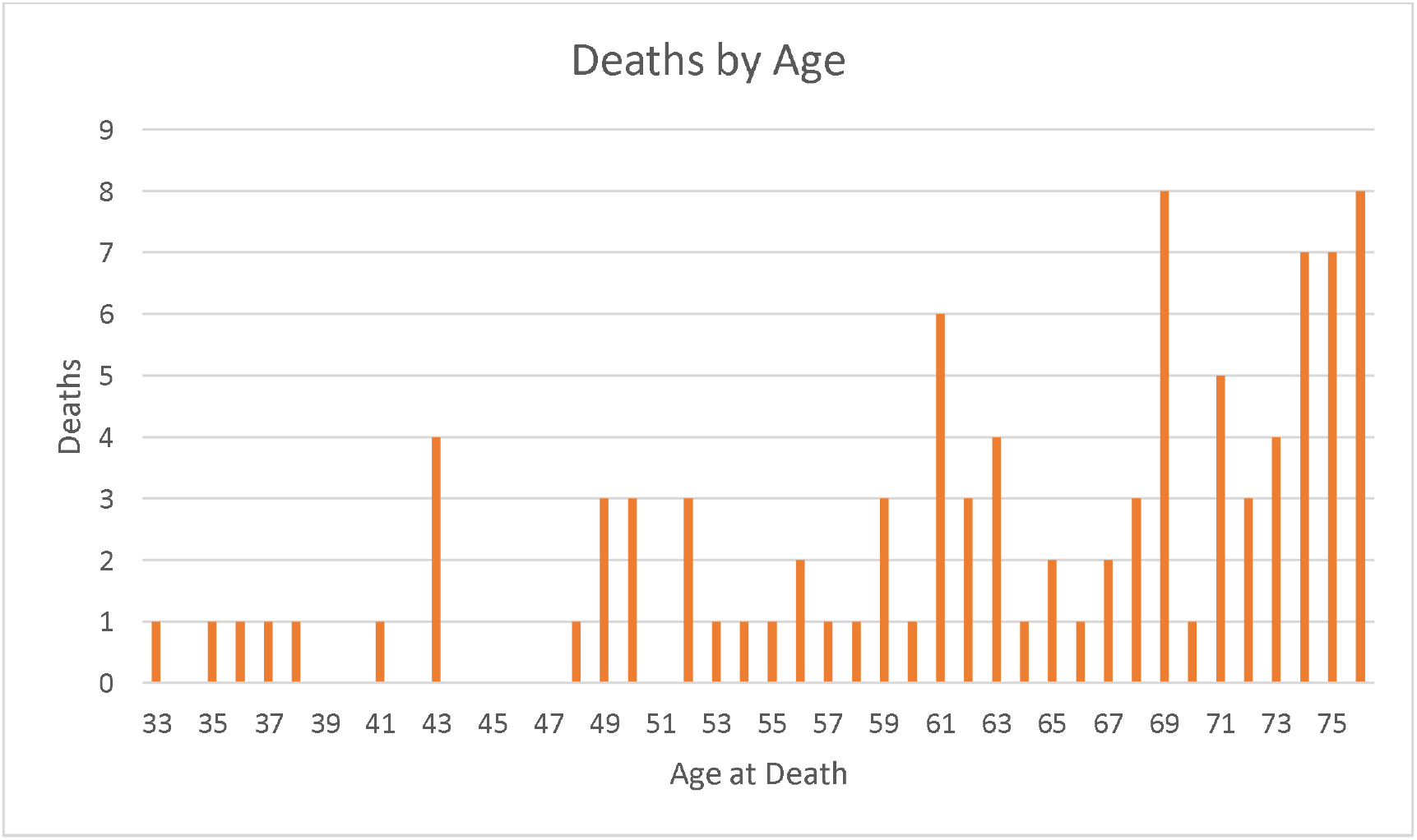

Comparing AFA67 and the NHIS sample for those aged 43 and greater, AFA67 has slightly lower mortality but the difference is not statistically significant (p=0.95). The added deaths in the AFA+12 cohort increase the age-specific death rates and reduce survival, but the difference between the two AFA67 scenarios is not statistically significant (p=0.34).

### Future projections

Table 2 lists the estimated deaths in the next five years, the number who will live to see their 100^th^ birthday, and average life expectancy at age 78 (the average age of living AFA67 graduates on September 1, 2022, was estimated to be 77.7 years). Noteworthy is the high level of uncertainty in the number living at age 100.

**Table 2.** Deaths in next 5 years, Number living at age 100, and Life expectancy at age 78.

|  | 130 known deaths | add 12 deaths |
| --- | --- | --- |
|  | (95% margin of error) |  |
| Deaths in next 5 years | 74<br>($\pm 28$) | 89<br>($\pm 32$) |
| Number living at 100 | 76<br>($\pm 48$) | 47<br>($\pm 38$) |
| Life expectancy at 78 | 13.8 years<br>($\pm 3.3$) | 11.9 years<br>($\pm 3.0$) |

Table 3 lists the estimated impact of self-reported health on life expectancy at age 78.

**Table 3.** Projected Life Expectancy at Age 78 by Level of Self-Reported Health.

| Self-Reported Health | (95% margin of error) |
| --- | --- |
| Excellent | 11.7<br>($\pm 1.3$) |
| Very Good | 10.9<br>($\pm 0.9$) |
| Good | 9.2<br>($\pm 0.8$) |
| Fair/Poor | 6.3<br>($\pm 0.9$) |

## DISCUSSION

The AFA67 survival experience closely matches that of contemporaneous NHWM college graduates in the US who were in excellent or very good health. This finding corroborates that of di Lego and Turra who found that education influenced longevity more than military training or stringent recruitment screening.^11^ Adding 12 deaths (a 9.2% increase in the number of deaths) does not make a statistically significant survival difference. However, the point estimate for life expectancy at age 78 for AFA67+12 is 1.9 years less. For those of us who have reached or are nearing age 80, that difference is not inconsequential.

We recognize that the AFA67 life expectancy in Table 2, 13.8 years, exceeds that of those in excellent health in Table 3, 11.7 years. These point estimates do not differ significantly and both estimates are based on relatively small sample sizes. More research is needed on the impact of education on mortality in the elderly. Furthermore, these estimates are greater than the overall NHWM life expectancies in the United States at age 78 in 2019 and 2020, 9.5 and 8.9 years respectively. The decrease from 2019 to 2020 reflects the impact of the COVID pandemic. The future impact of COVID and other potential mortality shocks on AFA67 and the US population is unknown.

Looking ahead, 46 to 102 (74 + 28) of the 386 living members of AFA67 on September 1, 2022, can reasonably be expected to die by August 31, 2027. In the 2019-2020 NHIS surveys the self-reported health proportions for NHWM college graduates aged 76-79 were excellent 19.5%, very good 39.7%, good 27.1%, and fair or poor 13.6%. If 13.6% of the living graduates were in fair or poor health, that equates to 53 of the 386 graduates assumed to be living in September 2022. Based on the monotonic decrease in life expectancy with self-reported health shown in Table 3, those in excellent health have a distinct survival advantage, 2.5 years compared to those in good health and 5.4 years more than those in fair or poor health. Thus, most of the decedents in the next five years will probably accrue from those who were in fair or poor health in September 2022. Between 28 and 124 (76 + 48) could live to age 100; this is a small sample, and the margins of error are large. It is important to recognize that our life expectancy, number of deaths in the next five years, and number living to age 100 estimates are based on the modeled age-specific mortality rates persisting into the future. The tenuous character of this assumption is exemplified by the increased mortality during the COVID pandemic.

These analyses have two important limitations, dates of birth, and dates of death. Although we believe actual birth dates would have little impact on the results, they would provide more confidence in the findings. Accurate death data are more critical. The NDI Plus is the central repository for deaths occurring in the United States since January 1979 and should be utilized. Moreover, although the Weibull distribution was used in the parametric survival modeling, other distributions, such as the Gompertz or gamma-Gompertz could be used. Crucially, we do not know the distribution of deaths among the elderly.^12^

These results should be considered exploratory and not confirmatory. They could lead to numerous other studies such as associations between longevity and active duty time, class rank at graduation, rank achieved after graduation, and the impact of flying status or occupational specialty. Two recent studies found increased cancer incidence and mortality in Air Force fighter aviators and military aviators in general.^13,14^ Although AFA67 graduates were included in the studies, it would be informative to focus on Air Force Academy graduates to examine multiple causes of death. For instance, the DoD aviator cancer study found markedly reduced lung cancer in military aviators. Reduced lung cancer mortality suggests low cigarette smoking rates, and these may translate to lower cardiovascular disease mortality.

AFA67 is too small a sample to provide definitive conclusions regarding causes of death. If the sample size were increased by tenfold, then given the applicability of normal theory, the margins of error could decrease by threefold. We view this study as an initial effort to investigate the methods and understand the level of effort needed to include more classes. Furthermore, examining the experiences of more service academy graduates is a distinct possibility—we envision a longevity research consortium involving all five service academies. Ideally, these studies would be performed by cadets and midshipmen with faculty guidance for the benefit of current and future graduates. Studies done in military settings would be advantageous because it should be easier to access military personnel records for more accurate dates of birth and some dates of death. Even more death data could be accessed through the Joint Department of Veterans Affairs and Department of Defense Suicide Data Repository.

## CONCLUSION

AFA67 experienced nearly fivefold excess mortality during the war in Southeast Asia. Thereafter, AFA67 survival closely matches that of contemporaneous non-Hispanic white male college graduates in the US who were in excellent health in their early twenties. There are numerous opportunities to extend this research that would be facilitated by involving the faculties and students at the five service academies.

## Data Availability

All data produced in the present study are available upon reasonable request to the author.

https://meps.ipums.org/meps-action/variables/group

## Acknowledgment

LS Dougherty and two other AFA67 graduates provided helpful review and constructive criticism of this manuscript.

