## Supplemental file for "United States Air Force Academy Class of 1967 Survival"

**Supplement**

Birthdates

As of September 1, 2022, there were 130 known decedents in AFA67. Using three sources (links at end), birthdates were obtained for 122 of the decedents. These birthdates are easily converted to ages.

In addition, we obtained birthdate data for a convenience sample of one squadron’s graduates. These included 15 living and 7 deceased who are also included in the 122 decedents mentioned above. Thus, there are birthdate data for a total of 137 of the 516 graduates who were commissioned leaving 379 without birthdate data. Following are summary age at graduation statistics for the 137 graduates with birthdates. Note the skewness to the right indicating the mean and median are greater than the mode.

| N | Minimum | Maximum | Mean | Std. Deviation | Skewness | Std. Error | Kurtosis | Std. Error |
| --- | --- | --- | --- | --- | --- | --- | --- | --- |
| 137 | 21.23 | 25.02 | 22.44 | 0.79 | 1.15 | 0.21 | 1.05 | 0.41 |

For the 379 graduates with missing birthdates, we generated simulated ages at graduation that conformed to the distribution of the known ages at graduation. Several distributions, including beta, chi-square, gamma, and skew-normal (SN) can have skewed probability density. We chose the SN as it provided a better fit. The SN distribution has an on-line random number generator at http://azzalini.stat.unipd.it/SN/sn-random.html.

The statistics below show the distributions’ similarity. The KS p-value is 0.55, indicating no significant difference in the distributions.

| Group | N | Minimum | Maximum | Mean | Std. Deviation | Skewness | Std. Error | Kurtosis | Std. Error |
| --- | --- | --- | --- | --- | --- | --- | --- | --- | --- |
| Known | 137 | 21.23 | 25.02 | 22.44 | 0.79 | 1.15 | 0.21 | 1.05 | 0.41 |
| Simulated | 379 | 21.33 | 25.51 | 22.44 | 0.70 | 1.15 | 0.13 | 1.48 | 0.25 |

Links to birthdate sources:

Findagrave.com (the best source) <https://www.findagrave.com/memorial>

a DoD database with Southeast Asia conflict decedents, <https://www.archives.gov/research/military/vietnam-war/casualty-statistics>

Social Security Death Index—no data before 1990 nor after 2012

<https://www.fold3.com/search?docQuery=(filters:!((type:general.title.id,values:!((label:Social+Security+Death+Index,value:%27830%27)))))>
